# Existential suffering as a motive for assisted suicide: difficulties, acceptability, management and roles from the perspectives of Swiss professionals

**DOI:** 10.1101/2022.03.21.22272665

**Authors:** Marie-Estelle Gaignard, Sophie Pautex, Samia Hurst

## Abstract

**Background:** Existential suffering is often a part of the requests for assisted suicide (AS). Its definitions have gained in clarity recently and refer to a distress arising from an inner realization that life has lost its meaning. There is however a lack of consensus on how to manage existential suffering, especially in a country where AS is legal and little is known about the difficulties faced by professionals confronted with these situations.

**Objectives:** To explore the perspectives of Swiss professionals involved in end-of-life care and assisted suicide on the management of existential suffering when it is part of AS requests, taking into account the question of roles, as well as on the difficulties they encounter along the way and their views on the acceptability of existential suffering as a motive for AS.

**Methods:** A qualitative study based on face-to-face interviews was performed among twenty-six participants from the fields of palliative and primary care as well as from EXIT right-to-die organization. A semi-structured interview guide exploring four themes was used. Elements from the grounded theory approach were applied.

**Results:** Almost all participants reported experiencing difficulties when facing existential suffering. Two-thirds considered existential suffering as a justifiable reason for requesting AS. Concerning the management of existential suffering, participants referred to the notion of being present, respect, explore the suffering, give meaning, working together, psychological support, spiritual support, relieve physical symptoms and palliative sedation.

**Conclusion:** This study offers a unique opportunity to reflect on what are desirable responses to existential suffering when it is part of AS requests. Existential suffering is plural and certainly implies a multiplicity of responses as well. These situations remain however difficult and controversial according to Swiss professionals. Clinicians’ education should better address these issues and give them the tool to take care of patients with existential suffering.

## BACKGROUND

Existential suffering is often a part of the requests for assisted suicide (AS).(1–3) The acceptability of existential suffering as a motive for AS is still a complex and controversial issue that is regularly debated in the medical literature.(4) According to recent definition, existential suffering refers to a distress arising from an inner realization that life has lost its meaning.(5– 8) Everybody can have existential concerns at any time of life but there is evidence that for patients at the end of life, those can increase their wish to hasten death.(9) As reported in our precedent paper, existential suffering was interpreted by Swiss professionals confronted to AS as a life that wasn’t worth living any longer and/or a life that did not make sense anymore, for many different reasons such as physical decline and loneliness.(10) We found that in most cases existential suffering consisted of different, and sometimes compounded, losses of the dimensions of life.

A certain clinical clarity on what existential suffering is is being acquired lately which should hopefully improve the evaluation and management of this kind of suffering in the coming years. Several questions then arise: how should we concretely manage existential suffering? After having explored and proposed other alternatives, is AS an acceptable solution to existential suffering? For recall, Switzerland has one of the most liberal legislations on AS in the world. The law does not specify requirements in terms of suffering. According to the SAMS (Swiss Academy of Medical Sciences), which provides healthcare professionals with recommendations on ethics issues, the assisting person has to verify that five conditions are present, including that the symptoms of the patient’s disease and/or her functional impairments are a source of “intolerable suffering” and that “medically indicated treatment options and other types of assistance and support have been sought and have provided ineffective or are rejected as unacceptable by the patient”.(11) This lack of consensus on what these approaches are and on how to manage existential suffering often leads to a feeling of helplessness in physicians and other health care professionals.(12–15)

Other authors, mainly from the palliative care field, have already addressed this issue and have suggested the importance of being present for the patient, the use of psycho-existential approaches and the promotion of spiritual care.(6,16–21) A 2008 review (9) reported on eight “existentially-informed interventions” addressing existential concerns that have the potential to positively impact a person’s well-being and overall quality of life at the end of life. Dignity therapy is one of the studied interventions that has certainly gained in recognition recently.(22) Although promising, these approaches are still too rarely used systematically in our hospitals. Another new and promising therapeutic approach is the use of classic serotonergic psychedelics in the management of patients with refractory depression and/or anxiety at the end of life.(23–25) Indeed, to date, no pharmacological approach has shown a significant benefit in the management of existential suffering. Palliative sedation is also subject to controversies.(2,13) The European Association for Palliative Care (EAPC) stated in 2009 that “palliative sedation may be considered for severe non-physical symptoms such as refractory depression, anxiety, demoralization or existential distress”, after taking into account several considerations.(1,4,26)

Another poorly investigated issue is that of roles in the management of existential suffering. So far, a few studies have pointed out the importance of different stakeholders such as nurses who are at the front line and can thus better recognize and evaluate existential suffering.(18,27,28) Mental healthcare professionals, such as psychotherapists and psychiatrists, and members of religious communities have also been recognized as playing an important role in the management of existential suffering.(13)

To our best knowledge, this is the first study investigating the perspectives of professionals on the management of existential suffering when it is part of the request for AS, all the more in a country where AS is legal. This paper, which constitutes the second part of our study, focuses on how Swiss professionals involved in end-of-life care and assisted suicide view the management of existential suffering, taking into account the question of roles. This study also explores the difficulties they may encounter along the way and their views on the acceptability of existential suffering as a motive for AS.

## METHODS

This exploratory national study involved a full spectrum of persons engaged in end-of-life care and assisted suicide in Switzerland. A detailed description of the method was reported in our previous paper.(10) **Table 1** summarizes its main features. A qualitative design was chosen for this study because of the lack of insights on the subject. Sampling was purposive, data were collected from face-to-face interviews, and data saturation was obtained after completing 20 interviews. Elements from the grounded theory approach were used to develop a conceptual model derived from the data.

**Table 1.**
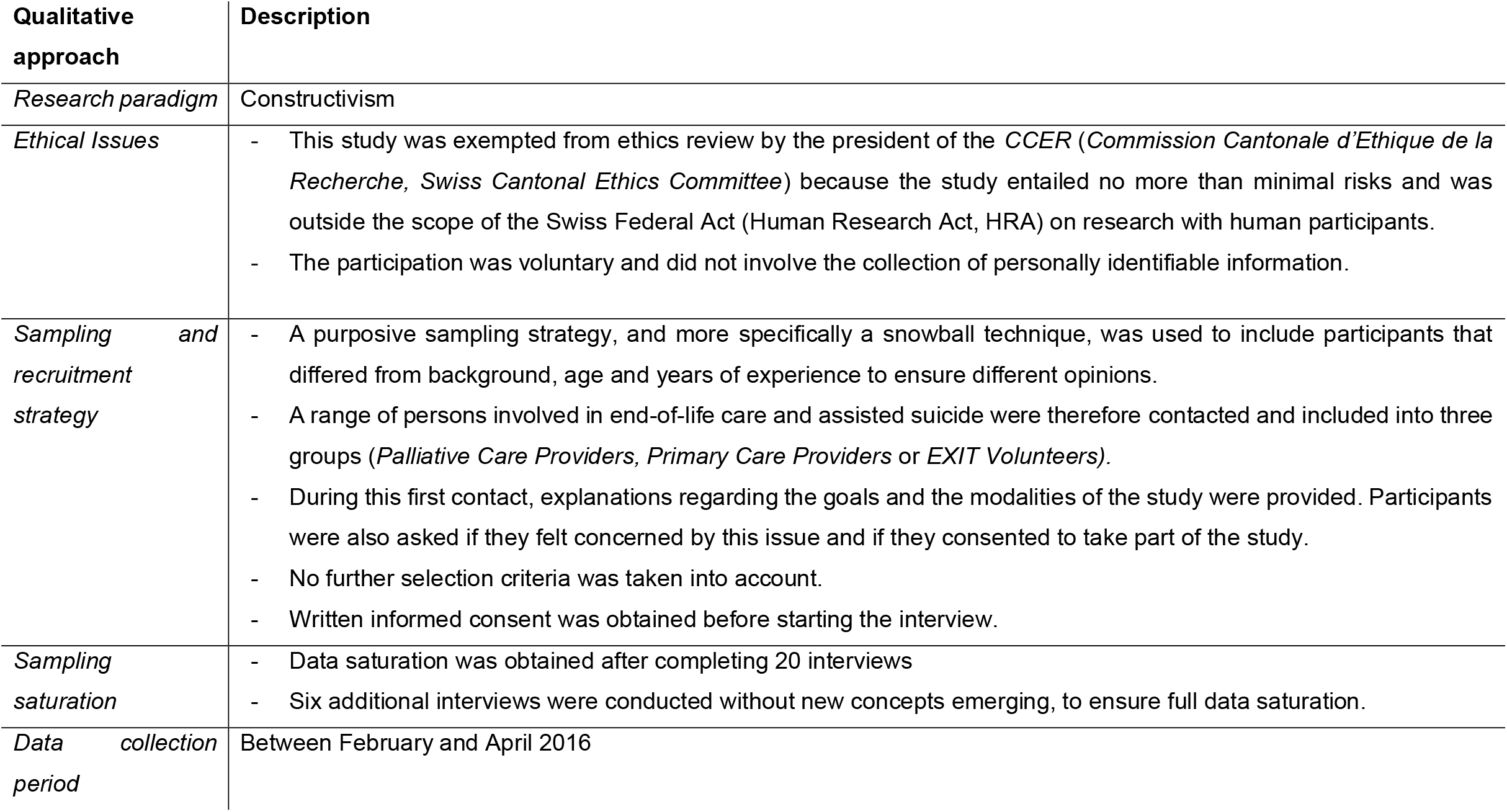
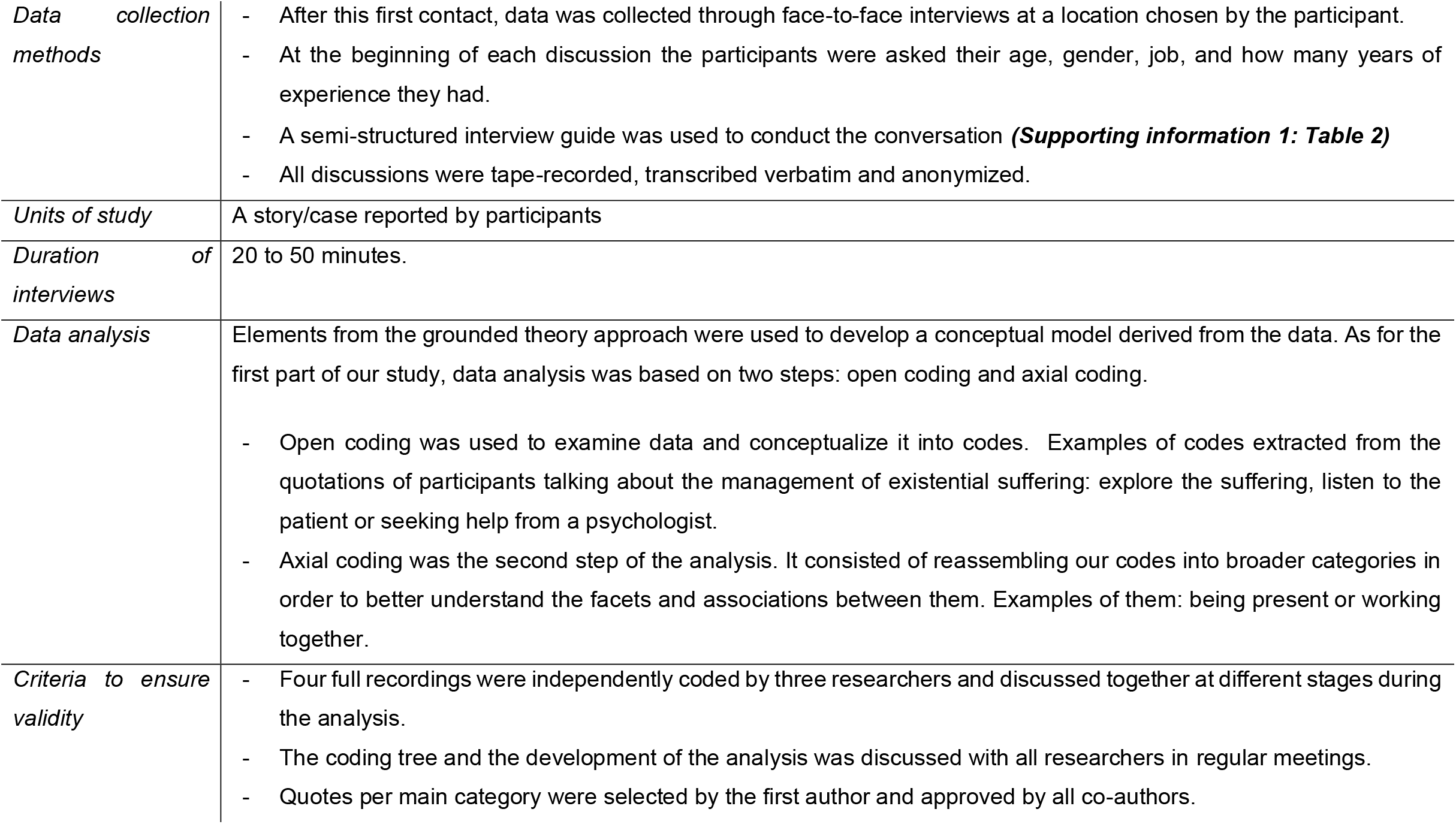
Design of the study.

In the following results, extracts of the quotations have been selected to illustrate ideas and concepts described by the participants. All transcripts were translated from the original French by the authors. People identification is labelled as follows: Pn (*Participant number*) / PallCP (*Palliative Care Providers*) or PrimCP (*Primary Care Providers*) or EV (*EXIT Volunteer)*.

## RESULTS

### Participants

As described in our previous paper (10), twenty-seven professionals were contacted between January and April 2016 to participate in this study. Only one did not answer our request, thus twenty-six people were interviewed. Participant characteristics are described in ***Supporting information 1: Table 1***. During one of the interviews, the recorder stopped after 10 minutes, without the investigator realizing it. We therefore decided to exclude this participant’s testimony from the second part of the analysis.

### Professionals’ perspectives on the management of existential suffering as a motive for requesting assisted suicide

#### 1. Difficulties when facing these kinds of requests

When professionals were asked if they encountered any difficulties when faced with these types of requests, 24 of them reported some kind of struggles. The coding of their statements resulted in two main types of difficulties: 1. Difficulties linked to a feeling of helplessness in the face of these situations and 2. Difficulties related to the conflict of values that these situations may cause. **Table 2** summarizes these findings and include quotes from participants. In general, most of them reported that these demands were challenging and that this could put them in difficulty. We found that, although most participants agreed that existential suffering is no less important than physical suffering, it remains somehow difficult to grasp and very controversial.

**Table 2.**
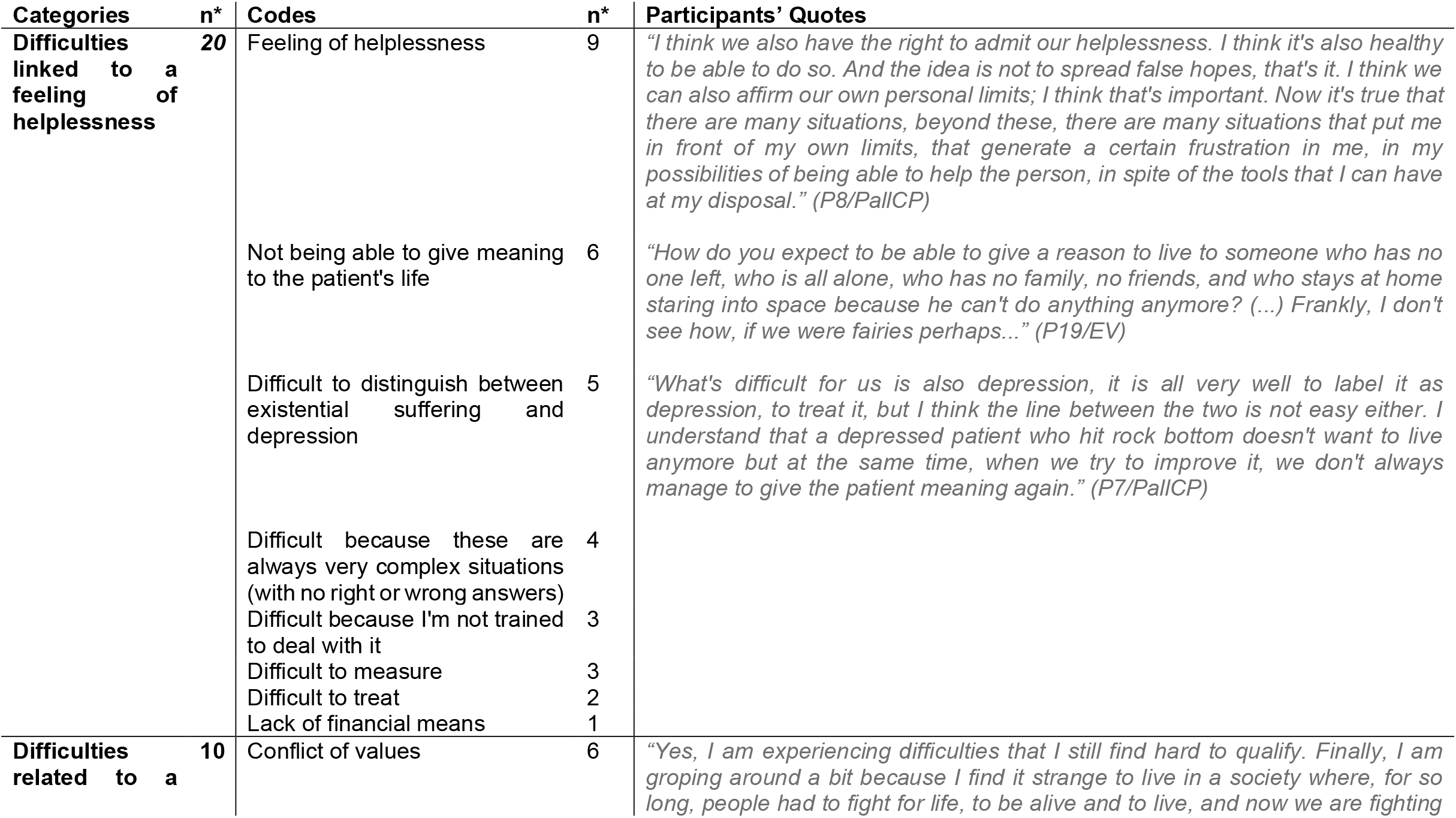
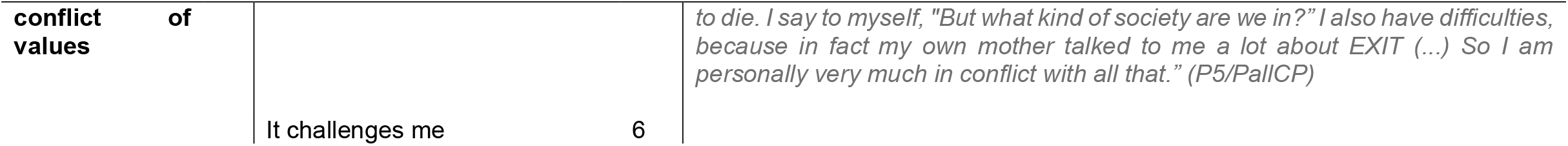
Difficulties encountered when facing existential suffering in AS requests. **n\***, number of participants who mentioned the category or code (N=25)

In spite of the above-described difficulties, 7 out of the 25 participants (of which 4 EXIT volunteers) said they were comfortable with these requests, even more so as we get older. Here is an illustration:

> *“The older I get, the more I am understanding I am of these demands. That’s what Mrs. X used to say: “She (another EXIT volunteer) is young, she believes that life is eternal. You’re old, you know you’re slowly going down so you’re aware of that. “” (P20/EV)*

#### 2. Acceptability of existential suffering as a motive for AS

A vast majority of the participants (17 out of 25) viewed existential suffering as a justifiable reason for requesting AS, especially as a last resort, after having exhausted all other alternatives. The main reason addressed was the fact that one cannot judge the suffering of others, because it is something so personal, intimate and because suffering cannot be limited to a disease status or a somatic cause. Here is an illustration by a palliative care provider:

> *“Yes, this is a fair reason for such a request. Well, she can of course ask because who am I to say she can’t? Then you have to question this request and then try to find out why, how and where it comes from, on what it is based. But the request is of course legitimate. Suicide is when the suffering becomes too great. All means are good as long as I can stop this suffering. And when I try a whole panoply of solutions that no longer work or that don’t work, well, that’s it, I’m going to look for others. And this (assisted suicide) is one of the solutions, of course. (P3/PallCP)*

Although existential suffering could be an acceptable reason for a request, it did not always mean for the participants that the request had to be granted. A primary care provider expresses this nuance:

> *“I think the patient’s request is legitimate. Totally legitimate in all cases. Now accessing it is another story. It’s really not the same thing. (…) It is always legitimate because that’s what they want and they have the right to want something because they are in a situation where they think this (AS) is necessary. That being said, in life there are a lot of things that we want but can’t have. And then the medical profession has to decide whether they want to become murderers or not*.*” (P24/PrimCP)*

Since the question in our interview concerned the acceptability of existential suffering in requesting AS, not in obtaining it, only a few participants went so far as to defend existential suffering as a justifiable reason for obtaining AS. On the contrary, 4 out of 25 participants considered that existential suffering was not a justifiable reason for requesting AS, as expressed here by an EXIT volunteer:

> *“I don’t think so, I don’t think so (that existential suffering is a justifiable reason for requesting AS). I can imagine that people are alone, abandoned, sad, but for me it is not a sufficient reason. I mean, there is no age for that. A young person aged 20 or 30 who is just fed up with it because life is too difficult and all, would it mean that we should consider AS for this kind of people? (*…*) Yes, there has to be some kind of physical suffering or a real loss of autonomy*.*” (P21/EV)*

Finally, 4 participants (out of 25) found it difficult to take a position on this issue, such as illustrated by this participant:

> *“To say what is right, not right, I can’t tell you. I’m a poor human being trying to get by. I think it’s a request that we can, that we must understand. If there is a request, we have to hear it*.*” (P14/PallCP)*

#### 3. Management of patients with existential suffering

On the basis of the professionals’ accounts, we identified 9 ways of managing existential suffering that were ranked according to the frequency of their occurrence: being present, respect, explore the suffering, give meaning, working together, psychological support, spiritual support, relieve physical symptoms and palliative sedation. A summary of these findings with quotes from participants can be found in **Table 3**.

**Table 3.**
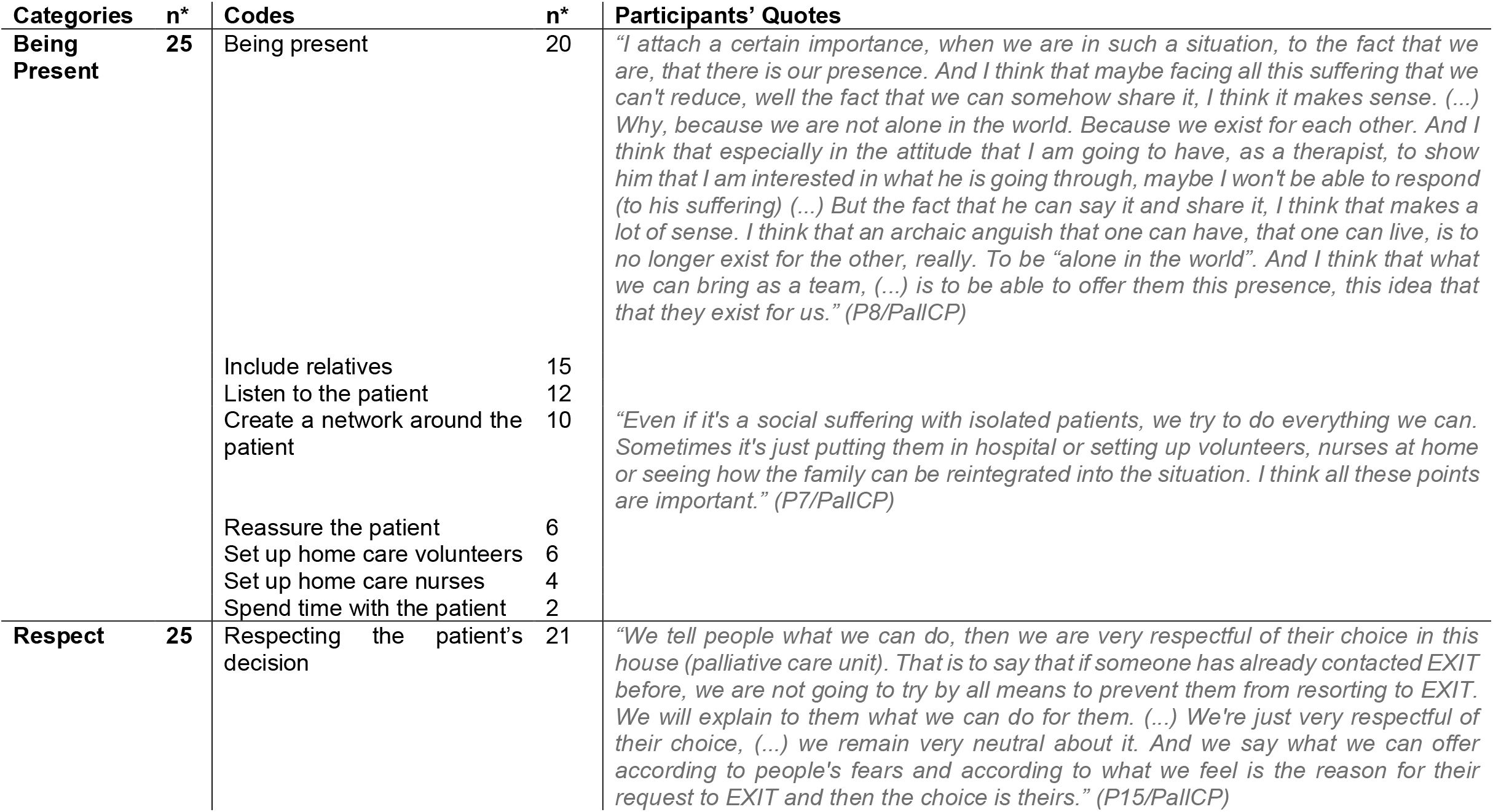
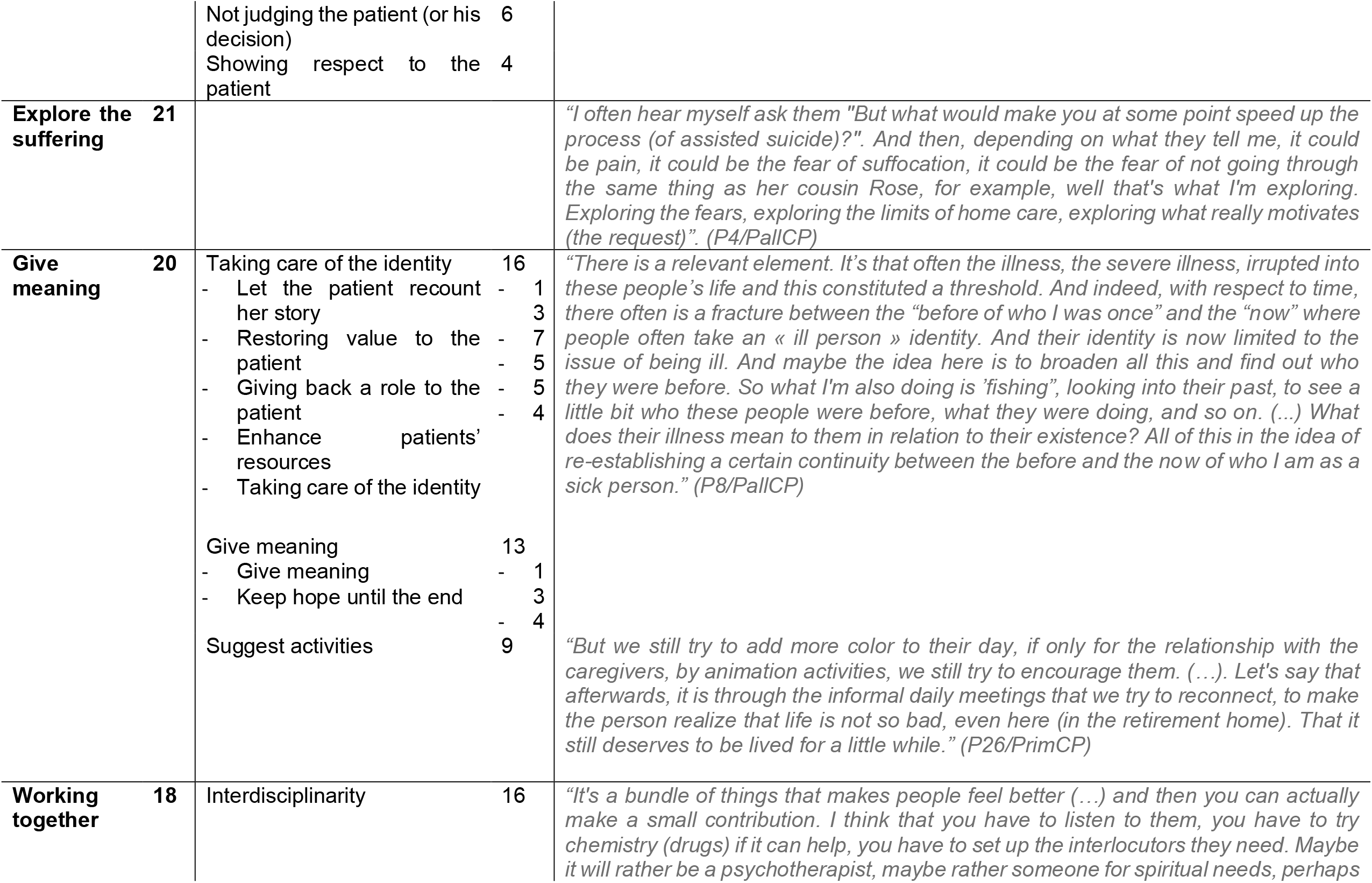
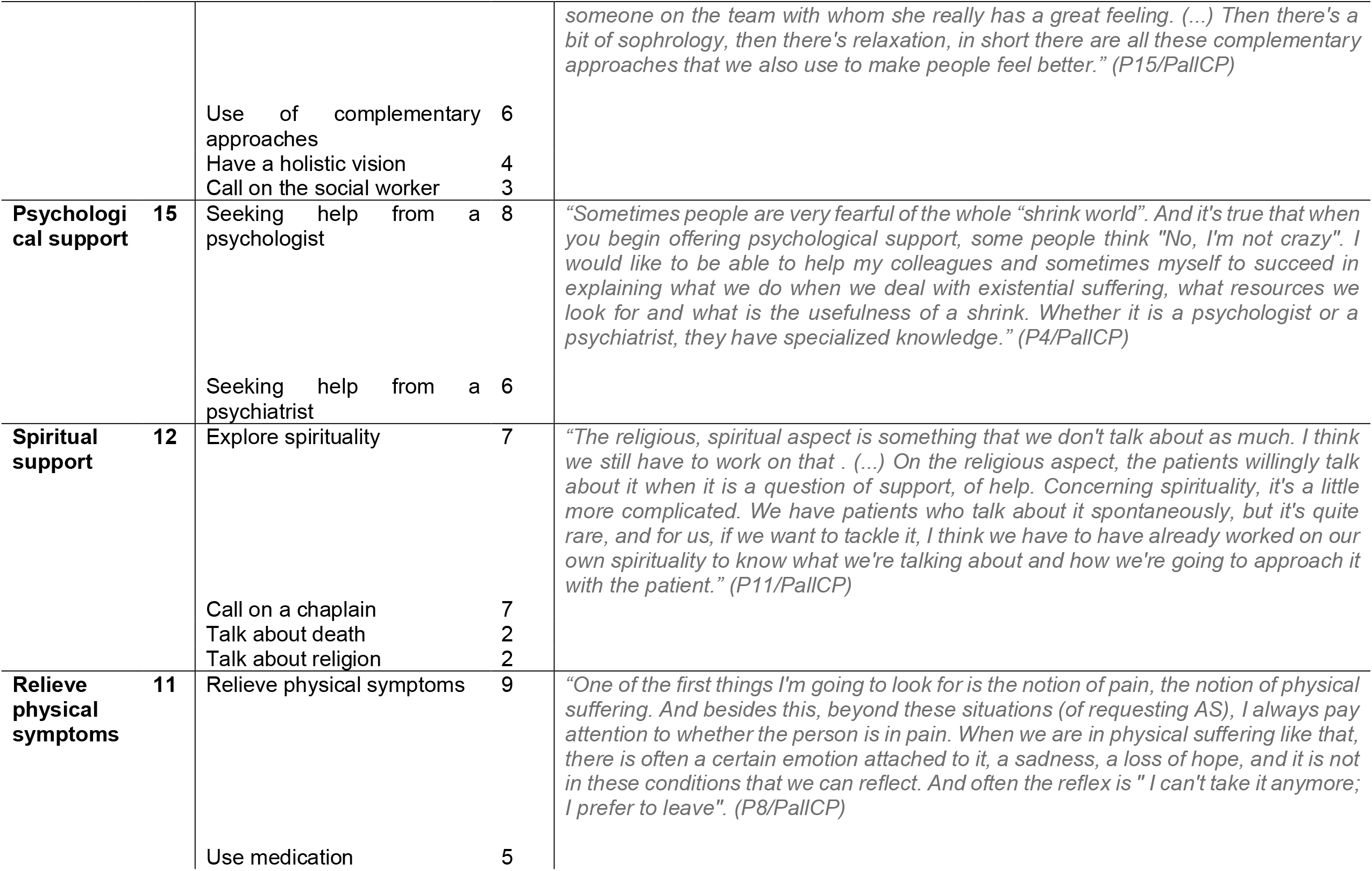
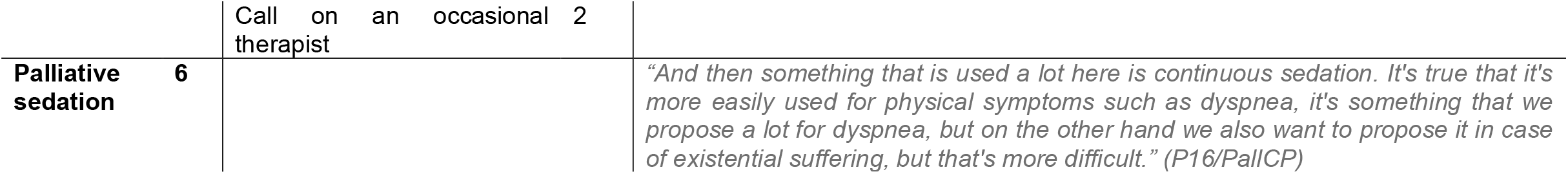
Management of existential suffering in AS requests. **n\***, number of participants who mentioned the category or code (N=25)

##### Being present

When asked about how existential suffering should be managed, all participants reported the importance of being present with your patient. Presence was seen as a way of creating this particular bond with the patient, allowing him to share his human experience, and his suffering. The presence as such was considered to be therapeutic, even if there were sometimes “no other solutions”. A few other codes were included in this category, such as including relatives, creating a network or listening, all highlighting how presence can be offered to patients.

##### Respect

All participants also referred to the importance of respecting the patient’s decision, not judging it, and showing respect to the patient. This category refers to an indispensable attitude to have, according to our participants, when supporting patients with existential suffering. Like the previous category, we can here note the emphasis placed by our participants on “being” rather than on “doing”.

##### Explore the suffering

A vast majority of the participants highlighted the importance of exploring the suffering of these patients, taking the time to understand their reasons for requesting AS. According to them, exploring the suffering was one of the essential first steps before being able to think of other alternatives.

##### Give meaning

20 out of the 25 participants reported the importance of giving meaning to their patients’ lives. This actually echoes their interpretations of existential suffering, as described in our previous paper: “a life that isn’t worth living any longer” and/or “a life that doesn’t make sense anymore”. According to these professionals, there are several ways to give meaning to their patients’ lives. The codes grouped in this category are: taking care of the identity, give meaning and suggest activities. Each of those includes sub-codes that are listed in **Table 3**. For many participants, a way to bring meaning back into someone’s life was to take care of the person’s identity, by helping her tell her story, by highlighting her identity and resources. While not referring to them by that name, these testimonials were reminiscent of narrative approaches that are used in existential psychotherapies.

##### Working together

The necessity of working together, of using “interdisciplinarity” was mentioned by more than two-thirds of the participants. It should be noted that this term of interdisciplinarity seemed sometimes vague. On the other hand, what was always emphasized was that it is never one person who holds the solution but that responses to existential suffering are plural and therefore emanate from many different people who have to collaborate.

##### Psychological support

When confronted to existential suffering, seeking help from a psychologist or a psychiatrist was suggested by many participants. According to our participants, mental health experts may have “better tools” to explore and to manage this type of suffering.

It should be noted that 3 participants thought the opposite: psychologists and psychiatrist would not be of great utility when managing existential suffering, as expressed by this participant:

> *“I think the psychologist is useless. Because what are these people going to believe? They will believe that we consider them sick people while I don’t think it’s a disease*.*” (P21/EV)*

##### Spiritual support

Half of the participants mentioned the usefulness of addressing people’s spiritual needs. The codes grouped in this category are: explore spirituality, call on a chaplain, talk about death and talk about religion. Their testimonials were not always clear on how to explore spirituality and many reported that it was not always easy to talk about it: because they didn’t have the skills, because it wasn’t “their job”, or because it was such an intimate subject and therefore difficult to discuss with patients.

##### Relieve physical symptoms

As expressed by this participant working as a psychologist in palliative care (see **Table 3**), 11 participants recalled the need to, first and foremost, properly manage physical suffering. Relieving physical symptoms was always seen as something we must have thought of before moving on to the next step.

##### Palliative sedation

A few participants working in palliative care mentioned the use of continuous sedation to alleviate existential suffering. As the quote in **Table 3** illustrates it, most said they were more comfortable when it was applied to the relief of physical symptoms but that this could happen, as a last resort and after having explored alternatives, in case of existential suffering, though recognizing that it was controversial.

#### 4. Whose role is it to manage existential suffering?

When participants were questioned about the roles around managing existential suffering, two types of responses emerged: 14 out of 25 thought that it is a shared responsibility among all health care professionals, pointing out the importance of interdisciplinarity. And among all of them, the most cited were, in order, physicians, psychologists, nurses and chaplains. This is illustrated by the testimony of a palliative care caregiver:

> *“Oh yes, I think that’s our role. It’s the role of an interdisciplinary team, well we’re all interested in it, it’s the role of all of us. And when we do interdisciplinary meetings, we try to put things in place at all levels, whether it’s at the level of the psychologist, the physician, the caregiver, the occupational therapist, the physiotherapist who can also suggest solutions, the social worker. There you have it, all the professionals revolving around the patient*.*” (P1/PallCP)*

This part of interview also allowed the participants to recall the importance of including patients’ relatives. Finally, among participants who thought existential suffering implies a shared responsibility, three of them even suggested that it was the responsibility of the entire society to better take care of these patients.

However, the rest of the participants (11 out of 25) expressed that this task, of managing existential suffering, was above all the prerogative of physicians, especially of general practitioners. We should here clarify that this task was seen more as that of a “conductor” who has to “coordinate” the care/management rather than do everything on his own.

As expressed here, it should be noted that among the six EXIT volunteers, only two considered that it was also their responsibility to manage existential suffering. The rest of the them thought it was not.

## DISCUSSION

Our results showed that almost all participants encountered difficulties when facing existential suffering, most of them related to a feeling of helplessness but also difficulties linked to the conflict of values that these situations can lead to. Regarding the acceptability of existential suffering in the context of an AS request, two-thirds of the participants considered it to be a justifiable reason for requesting AS. It should be noted that this did not necessarily mean that they agreed that this request should also be granted. Concerning the management of existential suffering, our participants expressed several key points and approaches to adopt when taking care of these patients. These include the notion of being present, showing respect, exploring the suffering, giving meaning, working together, psychological support, spiritual support, relief of physical symptoms and palliative sedation. As to the question of roles, the opinions of our participants were divided, with one party thinking that the responsibility should be shared among all health care professionals and the other thinking that it is mainly the prerogative of physicians.

Having already pointed out that existential suffering is plural, it is not surprising to find that the management of existential suffering has to be plural as well and involve an effective interdisciplinary care team. These results corroborate what previous authors suggested when aiming to relieve existential suffering.(17,19,20) This also fits with the organization of palliative care units in Switzerland, where interdisciplinarity is a concept and a method of operation that is now mostly well implemented.

As first steps in dealing with existential suffering, participants focused on crucial attitudes, rather than concrete approaches. Those include the notions of being present for your patient and showing her respect, qualities that are not necessarily taught in depth in medical schools. As Amonoo and colleagues (19) point out, “one of the privileges of being a clinician is being present for many of their patients’ significant moments – birth, death, life-altering illness, major accidents – but clinicians usually do not receive formal training to address the existential weight of these situations and it is not a clearly defined part of their role.” In this regard, several participants suggested that it was necessary to improve the training of physicians, to give them the tools to better listen and support patients with existential suffering. The hidden potentials behind a therapeutic presence have been well described by Covington (27) who defined “caring presence” as a “way of being—of deeply connecting—with another in a relationship” thus providing a “safe space for the patient to share suffering and find meaning in the illness experience”.

Also of interest, we found that all participants referred to themes related to existential psychotherapies, like meaning-centered approach or dignity psychotherapy, although they did not refer to them by this label. Those themes included the importance of giving meaning, let the patient recount her story or taking care of her identity. These approaches, drawing on the philosophy of existentialism pictured in particular by Victor Frankl and Irvin Yalom, surely represent a means of alleviating existential suffering by bolstering meaningful reflection at the end of life.(9,16) Unfortunately, most clinicians lack training in such approaches and this is certainly a path for improvement in the future. Amonoo and colleagues (19) also proposed a possible framework, utilizing existential themes, for addressing patients with existential suffering. This framework includes steps such as “take a narrative history to enhance perspective”, “help the patient actively enlist sources of resilience” and “help patients live in accordance with their core values”.

Only twelve out the twenty-five participants mentioned the value of exploring patients’ spiritual needs when facing existential suffering. This finding may seem rather low taking into consideration that this suffering is sometimes described as a spiritual one. These results, however, corroborate previous findings (29–31): patients at the end of life report that spiritual care is infrequently provided by their medical providers although they consider that religion and spirituality dimensions are important aspects of end-of-life care. This is also noteworthy because current palliative care guidelines include the importance of providing spiritual care.(21) A 2013-study asking the question “Why is spiritual care infrequent at the end of life ?” showed that lack of spiritual care training was the strongest predictor of spiritual care provision.(1,31) One can only speculate about the reasons underlying this finding but in secular hospitals, aiming to be open to all religious practices, one can perhaps expect this reserve towards spirituality, especially if it is viewed as mostly religious in nature. Future studies may be interesting to exploring this issue further.

The acceptability of palliative sedation for refractory existential suffering also raises controversial issues that have been explored in the literature recently.(4,12,13,32) A 2020 review (12) reports that “physicians do not hold clear views or agree if and when palliative sedation for existential suffering is appropriate” and that clinicians continue to be more favorable to palliative sedation for physical pain than for existential suffering. Ultimately, whether or not we are open to palliative sedation, it is our duty as clinicians, as stated by the SAMS (Swiss Academy of Medical Sciences) to have explored this suffering and to have proposed alternatives. Where to stop in the exploration of alternatives is however still an unanswered question. As with assisted suicide, the SAMS also sets out a normative framework for the use of palliative sedation in cases of existential suffering. These are based on the special considerations recognized by the EAPC (European Association for Palliative Care), in particular: the patient has to be in an advanced stage of a terminal illness, the refractoriness of his symptoms has been evaluated several times by clinicians skilled in psychological care, the evaluation has been made in the context of a multidisciplinary case conference, and repeated trials of respite sedation with intensive intermittent therapy have been performed.(26) Finally, regarding the question of roles, our study did not explore this issue in detail, but it did highlight the fact that all of us, as health care professionals, are concerned by this issue and that we all have a role to play in dealing with existential suffering. This surely includes physicians, who, according to almost half of our participants have an even greater role to play in the management of these patients. Unfortunately, studies on this topic show that patients are dissatisfied with the lack of concern of physicians and nurses for their existential needs.(33,34) This further reinforces the idea that clinicians’ education has to address these dimensions of care. Moreover, similar to psycho-existential approaches which do not necessarily need to be conducted by psychotherapists, providing spiritual care should not only be the prerogative of chaplains.(19,35)

As this is the first study investigating the perspectives of professionals on existential suffering, its acceptability, and its management in a country where AS is legal, it offers a unique opportunity to reflect on what are desirable responses to existential suffering when it is part of these requests. Existential suffering is plural and its management certainly implies a multiplicity of responses as well. Among these responses are the notions of being present, giving meaning, providing psychological and spiritual support, skills that are not, in modern medicine, necessarily assigned to physicians. As Kissane (20) points out, “the skills referred to herein are based on the physician as healer, listener, and doctor to the person rather than the symptom or disease”.

We hope that our findings will reinforce the work already underway to improve the training of clinicians in the exploration and treatment of existential suffering, as well as to perhaps more systematically implement psycho-existential approaches and spiritual care when confronted with these situations. All questions are far from being answered though and future research should focus on developing and implementing these approaches as well as to reflect on when it can be considered that the exploration of alternatives has been sufficient, implying that AS can become an acceptable alternative when facing existential suffering.

This study entails several limitations that have been described in our previous paper. Those include limitations related to the qualitative nature of the study and the fact that the sample size was small and only from the French-speaking part of Switzerland, leading to a lack of generalization of our results. This study also has the disadvantage of only focusing on the representations of professionals, and not on those of people requesting AS. However, we found that the solutions proposed here are close to what can be found in the previous literature as well as to the few studies that have reported patients’ wishes.(6,17–20) As mentioned in the results section, it is necessary to recall here the limitation of our interview question related to the acceptability of existential suffering as a motive for AS. Indeed, our question only involved the request and not the issue of granting the request. This may have overestimated the number of participants in agreement with the practice of AS in the case of existential suffering.

## CONCLUSION

This study brings to light that the management of existential suffering has to be plural and involve an effective interdisciplinary team. The approaches proposed here include the notion of being present, respect, explore the suffering, give meaning, working together, psychological support, spiritual support, relieve physical symptoms and palliative sedation. Our study also highlights that professionals confronted with these situations still encounter many difficulties and that their training needs to address these issues and give them the tool to better take care of patients with existential suffering. Reinforcing interdisciplinary and multiprofessional approaches is surely also key. We hope that this work will reinforce the efforts that are being made in this direction and that it will also stimulate the ongoing reflection on the acceptability of existential suffering as a reason for requesting AS.

## Data Availability

All data produced in the present work are contained in the manuscript

## ABBREVIATIONS

AS: assisted suicide
CCER: Commission Cantonale d’Ethique de la Recherche (Swiss Cantonal Ethics Committee)
EAPC: European Association for Palliative Care
EV: *EXIT* Volunteer
EXIT: name of a swiss right-to-die organization
Pn: Participant number
PallCP: Palliative Care Providers
PrimCP: Primary Care Providers
SAMS: Swiss Academy of Medical Sciences

## ACKNOWLEDGMENTS

The authors wish to show their gratitude to the participants in the study for their time and willingness to be interviewed. They would also like to thank their colleagues at the Institute of Ethics who provided insight on several occasions, as well as Claudia Ricci for reviewing the initial manuscript.

