## Supporting information for "Existential suffering as a motive for assisted suicide: difficulties, acceptability, management and roles from the perspectives of Swiss professionals"

### ES & AS – Supporting information 1

#### Table 1 – Number of participants and their demographics

**n*=**26

| | Palliative care providers (n*) | Volunteers from *EXIT* (n*) | Primary care providers (n*) |
|---|---|---|---|
| **People contacted** | 17 | 6 | 4 |
| **Participants** | 16 | 6 | 4 |
| **Gender** | | | |
| ⇨ **Female** | 10 | 3 | 4 |
| ⇨ **Male** | 6 | 3 | - |
| **Age range** | 43-62 | 49-80 | 52-63 |
| **Years of experience in their domain (range)** | 4-36 | 6-14 | 6-40 |

**Table 2 – Interview guide**

| TOPIC / THEME | MAIN QUESTIONS | FOLLOW-UP QUESTIONS |
|---|---|---|
| Experience with existential suffering and assisted suicide | Did you already receive requests for assisted suicide having existential suffering as a reason?<br><br>- if yes, can you tell me more about it?<br>- in your opinion, why were the reasons of this request “existential”?<br>- what finally happened to this person? | When you have been confronted to this existential suffering, did you explore it?<br><br>- if yes, how?<br>- if no, why? |
| Difficulties encountered | Do you feel any difficulties when you face these kinds of requests?<br><br>- if yes, which? | Do you think you have the tools to confront them?<br><br>- if yes, which?<br>- if no, what would you need? |
| Acceptability of existential suffering as a motive for assisted suicide | Do you think that existential suffering is a justifiable reason for requesting assisted suicide?<br><br>- why? | |
| Management and alternatives | 1) What should we propose to people requesting assisted suicide with a reason of existential suffering?<br><br>- which alternatives?<br><br>2) What would be your role?<br><br>- who else do you imagine having to take care of existential suffering when it’s part of the request for assisted suicide? | Do you think it is your role to explore existential suffering?<br><br>- if no, whose role, is it?<br><br>If the person doesn’t change his/her mind, how would you ensure that there is nothing else to do? |
